# Humoral response to the BBIBP-CorV vaccine over time in healthcare workers with or without exposure to SARS-CoV-2

**DOI:** 10.1101/2021.10.02.21264432

**Authors:** María Noel Badano, Florencia Sabbione, Irene Keitelman, Matias Pereson, Natalia Aloisi, Ana Colado, Victoria Ramos, Juan Manuel Ortiz Wilczyñski, Roberto Pozner, Luis Castillo, Georgina Wigdorovitz, María Marta E de Bracco, Susana Fink, Roberto Chuit, Patricia Baré

**Author notes:** **Corresponding author:** Patricia Baré. Postal address: J.A. Pacheco de Melo 3081, CABA 1425, Buenos Aires, Argentina. These authors contributed equally to this research (Florencia Sabbione, Irene Keitelman).

## Abstract

SARS-CoV-2-specific humoral response was analyzed over time in a group of healthcare workers with or without exposure to SARS-CoV-2, who underwent vaccination with BBIBP-CorV (Sinopharm) vaccine in Argentina.

Seroconversion rates in unexposed subjects after the first and second doses were 40% and 100%, respectively, showing a significant increase in antibody concentrations from dose 1 to dose 2 (*p*<0.0001).

The highest antibody concentrations were found in younger subjects and women, remaining significantly associated in a multivariable linear regression model (*p*=0.005).

A single dose of the BBIBP-CorV vaccine induced a strong antibody response in individuals with prior SARS-CoV-2infection, while a second dose did not increase this response. A sharp increase in antibody concentrations was observed following SARS-CoV-2 infection in those participants who became infected after the first and second doses (*p*=0.008).

Individuals with SARS-CoV-2 exposure prior to vaccination showed significantly higher anti-spike IgG antibody levels, at all-time points, than those not exposed (*p*<0.001). Higher antibody titers were induced by a single dose in previously SARS-CoV-2 infected individuals than those induced in naïve subjects by two doses of the vaccine (*p*<0.0001). Three months after the second dose both groups showed a decline in antibody levels, being more abrupt in unexposed subjects.

Overall, our results showed a trend towards lower antibody concentrations over time following BBIBP-CorV vaccination. Sex and age seem to influence the magnitude of the humoral response in unexposed subjects while the combination of exposure to SARS-CoV-2 plus vaccination, whatever the sequence of the events was, produced a sharp increase in antibody levels.

Evaluation of the humoral responses over time and the analysis of the induction and persistence of memory B and T cell responses, are needed to assess long-term immune protection induced by BBIBP-CorV vaccine.

## Introduction

In early 2020, the Beijing Institute of Biological Products developed an inactivated coronavirus vaccine called BBIBP-CorV (Sinopharm). This vaccine was authorized for emergency use on February 22^nd^ 2021 by the Ministry of Health of Argentina and on May 7^th^ 2021 by the World Health Organization (WHO), giving the approval for this vaccine to be rolled out globally with an efficacy estimate of 78.1%.

BBIBP-CorV is an inactivated vaccine consisting of virus particles that have been grown in culture and then inactivated to lose the ability to produce disease, while still stimulating an immune response. This vaccine was proven to be safe, immunogenic and effective in adults in phase 1/2 and phase 3 trials [1,2]. Recently it was also shown to be safe and immunogenic in people less than 18 years of age in a phase 1/2 trial [3]. Immunogenicity of the BBIBP-CorV vaccine was assessed by measuring neutralizing antibody responses, and no data are available about cellular memory response. Therefore, information on the induction and persistence of memory B and T cell responses, along with the evaluation of the humoral responses over time are needed to estimate the long-term immune protection against SARS-CoV-2 conferred by BBIBP-CorV vaccine.

In Argentina, BBIBP-CorV vaccine is one of the three most administered along with the Sputnik V (Gamaleya NRCEM) and ChAdOx1 nCoV-19 (University of Oxford/AstraZeneca) vaccines. Hence, in the current work we evaluated SARS-CoV-2-specific humoral response over time in a group of healthcare workers with or without exposure to SARS-CoV-2, who underwent vaccination with BBIBP-CorV vaccine in Argentina.

## Materials and methods

### Study design and participants

An ongoing longitudinal observational cohort study, started in February 2021 among healthcare workers from the Academia Nacional de Medicina, is being conducted to measure humoral and cellular responses over time after vaccination against COVID-19.

A group of 82 individuals who underwent vaccination with BBIBP-CorV vaccine was included. Among them, 66 subjects had no clinical history of SARS-CoV-2 infection, 8 individuals had a prior SARS-CoV-2 infection and 8 subjects got infected between doses or after vaccination: 4 more than 14 days after the first dose, 2 immediately after the second dose and 2 more than 14 days after the second dose. All SARS-CoV-2 exposed subjects had mild disease based on WHO classification [4].

As the BBIBP-CorV vaccine in Argentina was first authorized only for people less than 60 years old, most of our population was below this age, with the exception of 4 individuals (Table 1). With the aim of immunizing the largest number of people in the shortest time, second doses of vaccines were deferred in Argentina and, therefore, most of the individuals in our study group received their second dose 54 days after the first shot, instead of the original two-dose schedule 21 days apart.

**Table 1.** Characteristics of the study cohort.

| Table 1. Characteristics of the study cohort |  |  |  |  |
| --- | --- | --- | --- | --- |
|  | Unexposed<br>(n=66) | Prior SARS-CoV-2<br>infection (n=8) | SARS-CoV-2 infection<br>after dose 1 (n=4) | SARS-CoV-2 infection<br>after dose 2 (n=4) |
| **General characteristics** |  |  |  |  |
| Age (years) | 43 (26-80) | 44 (33-51) | 39 (26-48) | 44 (29-51) |
| Sex, *n* (%) |  |  |  |  |
| Female/Male | 50/16 (76/24) | 3/5 (38/62) | 4/0 (100/0) | 4/0 (100/0) |
| **SARS-CoV-2-related characteristics** |  |  |  |  |
| Mild cases, *n* | 0 | 8 | 4 | 4 |
| Time since infection to dose 1 (months) | NA | 6.5 (4-8) | NA | NA |
| SARS-CoV-2 infection after dose 1 (days) | NA | NA | 49 (37-49) | NA |
| SARS-CoV-2 infection after dose 2 (days) | NA | NA | NA | 12.5 (5-96) |

Values are expressed as median (range) or *n* (%). *NA* not applicable

Blood samples were drawn 21 to 30 days after the first (T1) and second (T2) doses, and three months after dose 2 (T3), to measure humoral response over time. For those subjects who had been infected by SARS-CoV-2 after the first and second doses, blood samples were also obtained 21 to 30 days after the SARS-CoV-2 infection. Plasma or serum samples were obtained immediately after centrifugation of the peripheral blood and stored in aliquots at -20°C until used.

This study was approved by the local Ethics Committee of the Academia Nacional de Medicina and written informed consent was obtained from each participant.

### SARS-CoV-2 antibody ELISA

SARS-CoV-2 spike-specific IgG antibodies were measured using the ELISA test COVIDAR according to the manufacturer’s instructions (Laboratorios Lemos S.R.L, Buenos Aires, Argentina) [5]. The plates of the assay are coated with a purified mixture of the spike protein and the receptor binding domain (RBD) of the SARS-CoV-2 virus. Applying the criteria used for the qualitative assay, by means of a cut-off value (the mean OD450 nm value of the Negative Control + 0.150) ± 10%, samples were considered as reactive, non-reactive or inconclusive. Antibody concentrations to SARS-CoV-2 spike protein expressed as International Units/mL (IU/mL) were determined by constructing a calibration curve with serial dilutions of the standard included in the immunoassay kit (400 IU/ml, reactive human serum adjusted to WHO First International Standard for human immunoglobulin against SARS-CoV-2, NIBSC Code 20/136, version 2.0 of 12/17 / 2020). Each sample was properly diluted to fit an OD 450 nm within the linear range of the calibration curve. Antibody concentrations were obtained by interpolating the OD 450 nm value for each sample into the calibration curve.

### Statistical analysis

Unpaired t test or Mann-Whitney test were used to assess differences in quantitative variables between two groups. Paired Wilcoxon signed-rank test was used to compare antibody levels before and after SARS-CoV-2 infection in subjects infected after the first and second doses. Categorical data were analyzed by the Fisher’s exact test. Multiple linear regression was performed to test potential associations of demographic variables (age and sex) and seroconversion after the first dose, with the magnitude of humoral immune response in unexposed subjects, by calculating the linear regression coefficient (β) with 95% confidence intervals (95% CI). For each time point studied, geometric mean concentrations (GMC) of anti-spike-specific antibodies levels with 95% CI were calculated. In all cases, a value of *p*< 0.05 was considered indicative of a significant difference. All data analyses were performed using the GraphPad 9.1.2 Prism software (GraphPad Software, San Diego, CA, USA).

## Results

### Antibody response in unexposed subjects

SARS-CoV-2-specific humoral response was analyzed in 66 SARS-CoV-2 naïve individuals who received the two doses of BBIBP-CorV vaccine. Median age of participants was 43 (26-80) and 76 % were women (Table 1). Twenty-six of 65 (40%) of the participants showed detectable antibody titers after the first dose (T1) with a GMC of 43.6 IU/ml (95% CI: 30.3-62.8). After the second dose (T2), all 60 tested subjects (100%) developed IgG anti-spike-specific antibodies (GMC: 377.0 IU/ml; 95% CI: 324.3-438.3), showing a significant increase in antibody titers from T1 to T2 (*p*<0.0001). Three months after the second shot (T3), the GMC dropped to 125.4 IU/ml (95% CI: 88.2-178.4) with antibody levels significantly lower compared to those present in T2 (*p*<0.0001), but still significantly higher than those corresponding to T1 (*p*<0.0001) (Figure 1A). Higher antibody values were observed both at T2 (*p*=0.07) and T3 (*p*=0.03), in those subjects who seroconverted after the first dose compared to those who did not.

**Fig. 1.**
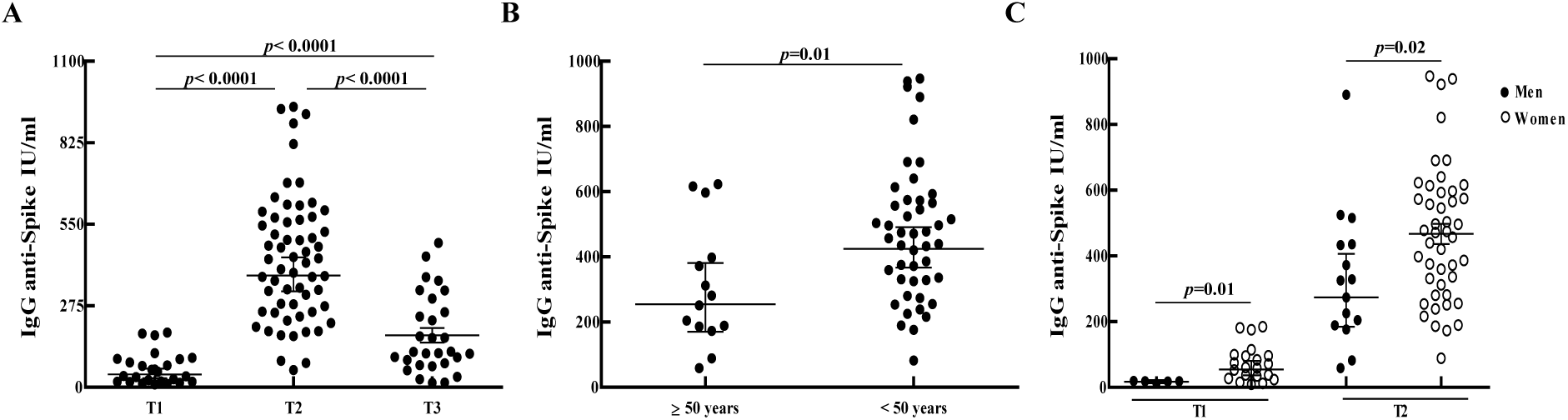
SARS-CoV-2-specific humoral response following vaccination with BBIBP-CorV in SARS-CoV-2 naïve individuals. (A) SARS-CoV-2 spike-specific IgG antibodies were measured 21 to 30 days after the first (T1) and second (T2) doses, and three months after dose 2 (T3). (B) IgG anti-spike antibody levels after dose 2 of BBIBP-CorV vaccine in subjects older and younger than 50 years. (C) anti-spike IgG antibody concentrations after the first and second doses of BBIBP-CorV vaccine in men and women. IgG anti-spike antibody concentrations are shown in International Units/mL (IU/mL). Geometric means with 95% confidence intervals are shown. *p* values were determined by Unpaired t test or Mann-Whitney test.

Lack of seroconversion after the first shot could not be associated with age or sex. However, after the second dose, subjects younger than 50 years old displayed significantly higher antibody concentrations than those over 50 (*p*=0.01) (Figure 1B). In addition, higher antibody levels were observed in women compared to men, after the first (*p*=0.01) and second doses (*p*=0.02) (Figure 1C). No differences were observed in the median ages between men and women (*p*=0.9). A multivariable linear regression analysis confirmed these result (*p*=0.005), showing that older age and male sex were significantly and inversely associated with anti-spike antibody levels at T2 (β1 (≥50 years):-158.5; 95% CI: -280.5-36.47; *p*=0.01; β2 (Male):-128; 95% CI: -247.2-8.784; *p*=0.03). As 76% in our study group were women, further analyses are needed to confirm this observation.

### SARS-CoV-2 infected subjects

Antibody responses were also analyzed in a group of 16 individuals who were infected with SARS-CoV-2 at different time points: before vaccination (n=8), after the 1^st^ dose (n=4) and after the 2^nd^ dose (n=4).

Antibody levels were analyzed longitudinally in 5 individuals with prior SARS-CoV-2 infection who got infected 4 to 8 months before receiving the vaccine (Table 1). All revealed seroconversion after the infection but one had no detectable antibodies at the time of receiving the first shot. The GM of IgG concentrations before vaccination (T0) was 203.2 IU/ml (95% CI: 42.9-962.4) and increased to 761.7 IU/ml (95% CI: 381.1-1522) after the first dose, showing detectable antibodies in all participants (*p*=0.06). Despite the small number of individuals to draw strong conclusions, no significant differences were observed in antibody levels between T1 and T2, reaching a GMC of 719.9 IU/ml (95% CI: 264.6-1959) at T2. Although there was a trend towards lower values three months after the second dose (GMC: 484.4 IU/ml; 95% CI: 147.3-1593), antibody levels did not significantly differ from those at T2 (Figure 2B).

Eight subjects became infected after the first or second dose of BBIBP-CorV vaccine. A sharp increase in antibody concentrations was seen after the infection (GMC: 2883 IU/ml; 95% CI: 1618-5137), being significantly higher compared to those present before the infection (GMC: 78.8 IU/ml; 95% CI: 14.7-423.7) (*p*=0.008) (Figure 2A). At T3, antibody concentrations remained high in this group of individuals (GMC: 1200 IU/ml; 95% CI: 491.3-2932), showing a trend towards lower values with similar levels to those observed at T3 in subjects infected with SARS-CoV-2 before vaccination (*p*=0.2). Further analyses at 6 and 12 months after vaccination would contribute to define the dynamics of the declining antibody titers in these particular groups (infection plus vaccination).

**Fig. 2.**
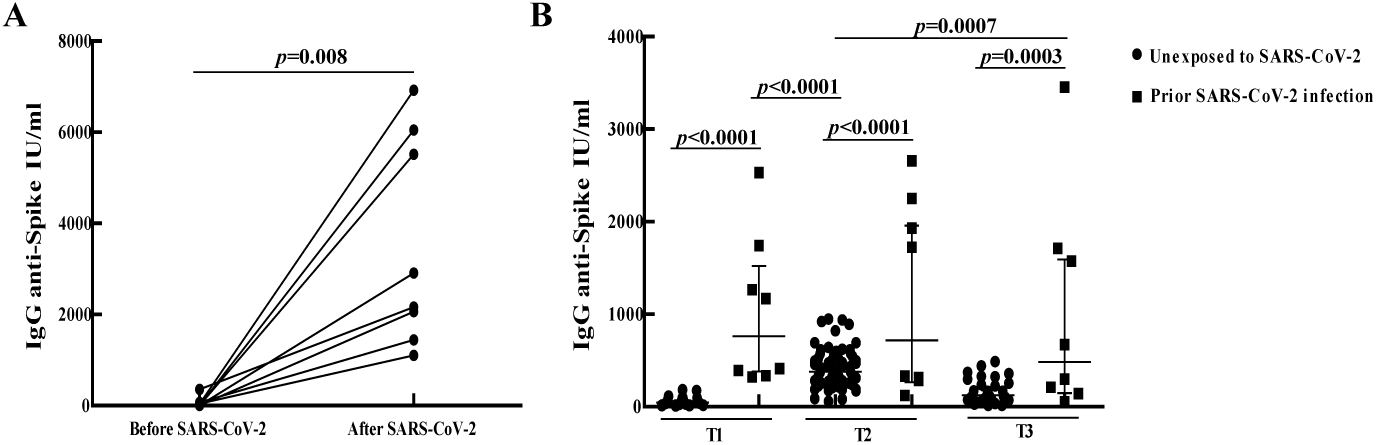
SARS-CoV-2-specific humoral response in BBIBP-CorV vaccinated subjects with exposure to SARS-CoV-2 virus. (A) IgG anti-spike antibody levels before and after SARS-CoV-2 infection in subjects infected after the first and second doses of BBIBP-CorV vaccine. (B) Comparison of anti-spike IgG antibody concentrations measured 21 to 30 days after the first (T1) and second (T2) doses, and three months after dose 2 (T3), between vaccinated subjects with and without prior SARS-CoV-2 infection. IgG anti-spike antibody concentrations are shown in International Units/mL (IU/mL). Geometric means with 95% confidence intervals are shown. *p* values were determined by the Wilcoxon matched-pairs signed-rank test (A) and the Unpaired t test or Mann-Whitney test (B).

### Comparison between unexposed and SARS-CoV-2 infected subjects

Antibody levels at different time points were compared between vaccinated subjects with and without prior SARS-CoV-2 infection. Individuals previously exposed to the virus displayed significantly higher concentrations of antibodies at T1 (*p*<0.0001), T2 (*p*<0.0001) and T3 (*p*=0.0003) compared to unexposed participants (Figure 2B), showing GMCs 17.5-, 1.9- and 3.9-fold higher at T1, T2 and T3, respectively. As it was reported for other vaccine formulations [6-9], antibody levels after one vaccine dose in previously infected participants were significantly higher (*p*<0.0001) than those reached after two doses in unexposed subjects (Figure 2B). Moreover, 3 months after the second dose individuals with prior SARS-CoV-2 infection still showed significantly (*p*=0.0007) higher antibody concentrations than those seen at T2 in SARS-CoV-2 naïve participants, when the peak in anti-spike-specific IgG antibody values occurred (Figure 2B).

## Discussion

In the current work, SARS-CoV-2-specific humoral response was analyzed in a group of unexposed and SARS-CoV-2 exposed subjects before or after the first or second dose of BBIBP-CorV vaccine. We observed that despite seroconversion rate after the first dose was low (40%) in SARS-CoV-2 naïve individuals, the 100% of participants developed antibody responses to BBIBP-CorV vaccine after the second dose. A decline in antibody levels was seen 3 months after dose 2, but remained higher compared to those present after the first dose. We also observed that age and sex influenced antibody concentrations reached after vaccination, since women and subjects under 50 years of age showed higher antibody levels than men and subjects over 50 years.

SARS-CoV-2 spike-specific IgG antibodies were also quantified in SARS-CoV-2 infected individuals. In agreement with other vaccine formulations [6-9], a single dose of BBIBP-CorV vaccine induces a strong antibody response in participants previously exposed to the virus, while a second dose appears not to increase this response. Three months after the second dose, antibody levels remained high but showing a decreasing trend, in this group. Measurement of SARS-CoV-2 spike-specific IgG antibodies in participants who became infected after the first or second vaccine doses, showed that antibody levels increased following SARS-CoV-2 infection and were significantly higher than those present before infection. Despite antibody levels remained high three months after dose 2, a trend towards lower antibody values was observed in this group. However, similar antibody concentrations were found at T3 between subjects exposed to SARS-CoV-2 before vaccination and those exposed after the first or second dose of BBIBP-CorV vaccine. These results suggest that exposure to SARS-CoV-2 increases antibody levels regardless of whether the infection occurs before, during or after vaccination.

Comparison between vaccinated subjects with and without prior SARS-CoV-2 infection showed that IgG anti-spike antibody levels were significantly higher in individuals with prior SARS-CoV-2 infection at all-time points studied. In line with other vaccine formulations [6-9], a single dose of BBIBP-CorV vaccine in previously SARS-CoV-2 infected individuals induces higher antibody concentrations than those seen in naïve subjects who received the two doses. Three months after the second dose, antibody levels in individuals with prior SARS-CoV-2 infection are still higher than those observed in SARS-CoV-2 naïve participants after the second shot, when the antibody values peaked. While both groups showed a decline in antibody levels at T3 (3 months after the second dose and 5 months after the first dose), this decrease was more abrupt in unexposed subjects. These results suggest that, as a consequence of their higher antibody responses following vaccination, individuals with prior SARS-CoV-2 infection would display detectable antibodies in blood for longer periods than unexposed subjects. As this is an ongoing study, IgG antibody titers will be measured at 6 and 12 months after vaccination to monitor the humoral response over time, in the different groups studied.

Immune response after vaccination is usually monitored by the measurement of antibody levels because is cheaper, easier and fast compared to immune techniques that detect and evaluate the functional capacities of immune memory cells. Therefore, correlates of protection after vaccination are generally determined by establishing a minimum threshold antibody level. So far, the threshold needed to confer protection against SARS-CoV-2 virus after COVID-19 vaccination is unknown. In the current work, in spite of 100% seroconversion rate achieved with a two-dose schedule of BBIBP-CorV vaccine, a decrease in antibody levels was seen over time. However, although antibodies wane over time following vaccination, this does not necessarily imply that immune memory response is lost and therefore, immune protection. In fact, we recently showed the presence of hepatitis A virus (HAV)-specific memory CD4^+^ T cell responses in 54% of pediatric individuals who had received a single dose of inactivated HAV vaccine 12 years before and lacked protective anti-HAV antibody levels (≥ 10 mIU/mL), suggesting that the presence of immune memory cells could be contributing to the absence of new HAV outbreaks in our country [10]. Thus, information about memory T and B cell responses mounted after BBIBP-CorV vaccine and its persistence over time is needed. Ongoing experiments to study SARS-CoV-2-specific memory cell responses in this cohort of BBIBP-CorV vaccinated individuals are being performed by our lab.

The seroconversion rates 14 days after the first and second doses of the BBIBP-CorV vaccine were reported to be 75% and 100%, respectively [1,2]. In the present work, seroconversion rates were 40% and 100%, 21-30 days after the first and second doses, respectively. This could be explained by the different methodologies used to evaluate seroconversion (binding versus neutralizing antibody assays) or different timing in seroconversion of the cohorts evaluated. It is important to emphasize that even though our group received the second dose more than 21 days apart, a seroconversion rate of 100% was achieved.

As reported, the magnitude of neutralizing antibodies induced by BBIBP-CorV vaccine was higher in individuals up to 60 years compared to those aged 60 years and older [1]. In agreement, we observed higher antibody concentrations after two doses of BBIBP-CorV vaccine in subjects younger than 50 years compared to those older than 50 years.

An association between the magnitude of the humoral response after BBIBP-CorV vaccination and sex has not been reported. In a recent work using SARS-CoV-2 proteins and peptides microarrays to analyze the humoral response elicited by the inactivated vaccine, a trend towards higher IgG anti-spike antibody signal was observed in women compared to men [11]. In the present study, women showed significantly higher antibody levels compared to men, after the first and second doses. These discrepancies could be explained by the difference in sample size and proportion of women tested and/or the different techniques used to analyze SARS-CoV-2 humoral responses. As the majority of individuals in our population were women, these observations need to be confirmed

A higher seroconversion rate after the first dose and higher levels of anti-spike-specific antibodies were found using the same ELISA test (COVIDAR-IgG) when evaluating the humoral response induced following Sputnik V vaccination in unexposed subjects [8, and our unpublished results]. However, Sputnik V vaccine only targets the spike protein, while inactivated virus vaccines could theoretically target spike plus other viral proteins. Therefore, it is possible that the humoral response induced by BBIBP-CorV vaccine is more diverse and targets other antigens of the SARS-CoV-2. Indeed, a recent work showed that higher antibody responses to the non-structural proteins NSP7, NSP8 and the RNA-dependent RNA polymerase (RdRp), which are part of the viral replication complex, are induced in healthy individuals following vaccination with BBIBP-CorV when compared to those observed in COVID-19 recovered subjects [11]. However, it could also be possible that the Sputnik V vaccine induces a stronger humoral response against the SARS-CoV-2 spike protein than that induced by the BBIBP-CorV vaccine.

This study has some limitations. Most of the unexposed subjects have unknown serostatus prior to vaccination. However, the low seroconversion rate after the first dose and the abrupt decline in antibody levels 3 months after the second dose, strongly suggest that they were seronegative at baseline. Antibody neutralization assays were not performed and therefore should be done to estimate long-term protection after BBIBP-CorV vaccine. Despite the low number of SARS-CoV-2 exposed subjects to draw strong conclusions, the same tendency for other vaccine platforms was observed, showing that robust antibody responses are elicited after one shot of BBIBP-CorV vaccine in individuals with prior infection.

In summary, in the present study we quantified anti-spike antibodies over time after vaccination with BBIBP-CorV vaccine in terms of IU/ml, allowing assays from different laboratories to be compared. Despite the 100% seroconversion rate achieved after the second dose, a decline in anti-spike antibody levels was seen 3 months after dose 2. Several factors like sex, age and exposure to SARS-CoV-2 seem to influence the magnitude of this humoral response, as the highest antibody concentrations in unexposed subjects were seen in younger subjects and women, and the combination of exposure to SARS-CoV-2 virus plus vaccination, whatever the sequence of the events is, produces a sharp increase in antibody levels.

Overall, our results suggest that the evaluation of the humoral response over time, through the measurement of the antibody levels against spike and other SARS-CoV-2 proteins along with the neutralization titers, and the analysis of the induction and persistence of memory B and T cell responses, are necessary to assess long-term immune protection induced by BBIBP-CorV vaccine.

## Data Availability

not applicable

## Declaration of Competing Interest

The authors declare that they have no known competing financial interests or personal relationships that could have appeared to influence the work reported in this paper.

## Declaration of Author Contributions

Conceptualization, MNB and PB, general coordination, MNB, FS, IK, RC and PB, collection of blood samples and clinical data, MNB, FS, IK, MP, NA, AC, VR, JMOW, RP, LC, GW and PB, determination of IgG anti-spike antibody concentrations, MNB, MP, NA, and PB, data curation and analysis, MNB, MMEB, SF, RC and PB, writing – original draft, MNB and PB, Writing – review & editing, MNB, FS, IK, MP, NA, AC, VR, JMOW, RP, LC, GW, MMEB, SF, Rc and PB. All authors read and approved the final manuscript.

## Acknowledgments

The authors thank Fabiana Alberto, Santiago Castera, Bárbara Giménez and Macarena Asensio for collaboration with blood draws. The authors are grateful to the COVIDAR group for providing ELISA antibody tests (COVIDAR-IgG). Some aspects of this work could not have been fulfilled without the generous contribution of the Fundación René Baron, IIHEMA, IIE and Academia Nacional de Medicina who provide financial support to our ongoing research.

## Notes

### Competing Interest Statement

The authors have declared no competing interest.

